# The impact of COVID-19 on population cancer screening programs in Australia: modelled evaluations for breast, bowel and cervical cancer

**DOI:** 10.1101/2022.09.11.22279831

**Authors:** Carolyn Nickson, Megan A Smith, Eleonora Feletto, Louiza S Velentzis, Kate Broun, Sabine Deij, Paul Grogan, Michaela Hall, Emily He, D James St John, Jie-Bin Lew, Pietro Procopio, Kate T. Simms, Joachim Worthington, G Bruce Mann, Karen Canfell

## Abstract

**Background:** Australia introduced COVID-19 infection prevention and control measures in early 2020. To help prepare health services the Australian Government Department of Health commissioned a modelled evaluation of the impact of disruptions to population breast, bowel and cervical cancer screening programs on cancer outcomes and cancer services.

**Methods:** We used the *Policy1* modelling platforms to estimate outcomes for potential disruptions to cancer screening participation, covering periods of 3, 6, 9 and 12 months. We estimated missed screens, clinical outcomes (cancer incidence, tumour staging), and various diagnostic service impacts.

**Results:** We estimated that a 12-month screening disruption would reduce breast cancer diagnoses (9.3% population-level reduction over 2020-2021) and colorectal cancer (up to 12·1% reduction over 2020-21), and increase cervical cancer diagnoses (up to 3·6% over 2020-2022), with upstaging expected for these cancer types.

**Conclusions:** Findings illustrate that maintaining screening participation is critical to sustaining a reduced cancer burden. We provide program-specific insights into which outcomes are expected to change, when changes are likely to become apparent, and likely downstream impacts. This evaluation provided evidence to guide decision-making for screening programs, and emphasises the ongoing benefits of maintaining screening in the face of potential future disruptions.

**Funding:** Australian Government Department of Health

## Introduction

Population cancer screening programs aim to reduce cancer-specific mortality, complications and side-effects associated with treatment of advanced stage neoplasms and, in the case of cervical and colorectal screening, reduce the incidence, illness and mortality of cancers through the detection and treatment of precancerous abnormalities.^1-3^ Breast, cervical and colorectal cancer screening programs reduce cancer-specific deaths, with mortality benefits outweighing the risks associated with screening.^1-3^

Australia established national screening programs for breast and cervical cancer in 1991 and colorectal cancer in 2006.^4-6^ Following the World Health Organisation’s declaration of the COVID-19 pandemic, in March 2020, Australia introduced strict national and state-level infection control measures including physical and social restrictions affecting health services as well as business, transport, and public gatherings. These restrictions proved highly effective for short-term national pandemic control, although significant outbreaks followed through returning travellers and hotel quarantine programs,^7^ subsequent new COVID variants ^8^ and ongoing variation in control measures.^9^ It was not known to what extent restrictions would impact cancer screening participation, nor the implications of pauses in delivering screening programs.

Modelled evaluations of COVID-impact scenarios have been undertaken by different countries to support their cancer screening programs as they respond to the pandemic. ^10-12^ In parallel, collaborations within the global modelling community have been established, such as the COVID-19 and Cancer Global Modelling Consortium, to collectively conduct evaluations to support cancer control during and after the COVID-19 pandemic.^13^ In early 2020, we were commissioned by the Australian Government Department of Health to undertake a rapid-response modelled evaluation of the health impacts of potential widespread disruptions to population breast, bowel and cervical screening. This was the first time a country-level analysis across all screening programs had been performed.

We report here on key findings from this evaluation, and discuss how they relate to subsequent observed disruptions to screening.

## Methods

We used simulation modelling to estimate outcomes for various potential disruptions to cancer screening services. Modelled scenarios were developed after consultation with the Australian Government Department of Health and input from administrators of the national screening programs, and were designed to be within the capabilities of our comprehensive and validated *Policy1* modelling platform (www.policy1.org).^14-15^ Scenarios, reported outcomes, and calendar periods varied between screening programs due to differences in cancer natural histories, existing screening program protocols, and stakeholder requests and advice.

### Modelling platform

To simulate scenarios and estimate projected outcomes, we used *Policy1* to evaluate the clinical benefits and harms of a wide range of screening protocols, new technologies and risk-based approaches aiming to optimise population cancer screening programs. Further details are provided in Supplementary Materials.

## Scenario selection

Scenarios were specified according to screening disruption duration (broadly 3, 6, 9 or 12 months) and degree (complete pauses or reduced throughput) (Table 1).

**Table 1:** Scenarios modelled for each of the three screening programs and key outcomes reported. Each scenario is compared to status quo outcomes, which incorporate various population projections based on pre-COVID observed data.

| Screening program | Disruption scenario by period |  |  |  | Outcomes |  |  |
| --- | --- | --- | --- | --- | --- | --- | --- |
|  | 12 months | 9 months | 6 months | 3 months | Cancer diagnoses | Delayed diagnoses/cancer staging | Screening episodes |
| **Breast screening (females, 50-74 years)<sup>1</sup>** | 12-month pause assuming gradual recovery<sup>1</sup> | N/A | 6 month-pause assuming gradual recovery<sup>1</sup> | 3-month pause assuming gradual recovery<sup>1</sup> | Invasive breast cancers (2020-2021) | Interval cancer rates, tumour size, grade and nodal involvement at diagnosis (2020-2021) | Screening episodes (1 April 2020- 31 March 2021) |
| **Bowel screening (persons, 50-74 years)<sup>2</sup>** | 12-month pause | N/A | 6-month pause | 3-month pause | CRC cases (2020-2021; 2020-2060) | Adenomas missed or delayed (2020-2021)<br>CRC cases detected at a later stage (2020-2060) | Number of iFOBT screening kits returned (2020-2021) |
| **Cervical screening (females, 25-74 years)<sup>3</sup>** | 12-month disruption assuming a 95% reduction in primary screening attendance | 9-month disruption assuming a 75% reduction in primary screening attendance | 6-month disruption assuming a 50% reduction in primary screening attendance | N/A | Additional cervical cancers (2020-2022) | Cervical cancers detected at a later stage (2020-2022) | Number of women screening (2020-2022) |

Breast screening: status quo: biennial mammography for women 50-74 years old; invitation letter sent at age 50 years; 55% participation rate. All scenarios assume no throughput during the pause period then a gradual recovery of screening throughput, reaching status quo rates by the seventh month after screening resumption. Breast screening outcomes are reported for women aged 50-74 at any time during the reporting period. Two-year reporting periods align with routine reporting of BreastScreen outcomes due to the program being mostly biennial.<sup>4</sup> Breast program recall rates describe mammographic screening episodes referred for further assessment, and false positive recall rates describe recalled screens with a benign final outcome after assessment. Screening program sensitivity describes the screen-detected cancers as a proportion of screen-detected cancers and interval cancers (further detail in Supplementary Materials).
Bowel screening: status quo: biennial iFORBT screening of men and women 50-74 year old; invitation letter with test sent at age 50 years; participation rate 43.5%. No throughput is assumed during the pause period. Bowel screening outcomes are reported for individuals age-eligible (aged 50-74 years) for screening in either 2020 or 2021 i.e., individuals born in 1945-1971, with reporting periods selected to indicate both the immediate effect and the lifetime effect of the eligible cohort. Outcomes describe the expected number of incident colorectal cancers in the screening cohort only.
Cervical screening: status quo: 5-yearly HPV screening of women 25-74 years old; invitation letter sent at age 25 years; participation rate at 46%. Primary screening tests describe women attending for a primary screening test. Scenarios assumes women who miss screening attend over 2021-2022, no disruption to surveillance or colposcopy visits, and no changes to rates of women presenting with symptoms. Cervical screening outcomes are reported for women age-eligible (aged 25-74 years) for screening at any time during 2020-2022, reporting the period 2020-2022 because all women who missed screening in 2020 were assumed to re-attend by 2022, and because 2022 aligns with the last year in the first 5-year-round of primary HPV screening.

### Breast screening

The BreastScreen Australia (BSA) program offers biennial (and some annual) mammographic screening to women from the age of 40 years, targeted to women aged 50-74 years. ^4^ The program is administered by the eight state and territory governments under a national agreement. All abnormal screens are assessed within the program. Prior to COVID-19, participation by women aged 50-74 was approximately 55%, with 51% of national breast cancers detected through BreastScreen, and around 60-84% of clients screened on schedule (first to third and subsequent round).^4^ Using the Policy1-Breast model, we evaluated a 3-, 6-, or 12-month national-level pause (applied as an averaging of annual rates), commencing 1 April 2020, assuming a gradual restoration of service throughput in the first 6 months, reaching prior throughput by the seventh month after screening resumption. Opportunistic or risk-based screening outside the program was assumed to be reduced by 50% during program pause (due to COVID-19 social distancing requirements), reaching 100% on BreastScreen service resumption (Figure supplement 1 and 2). In periods of reduced throughput, available screens were prioritised to women in the target age range of 50-74 years and to clients most overdue for screening. For this evaluation, Policy1-Breast was revised to include a prioritisation module (supplementary file 1A) so that it could be applied to capacity-driven recovery scenarios. With this revision, individual clients within the simulation could be channelled into available screening appointments prioritised according to different factors, such as age or screening round.

Additional information on the Policy1-Breast model can be found at www.policy1.org/models/breast.

### Bowel screening

The National Bowel Cancer Screening Program (NBCSP) provides biennial primary screening, using an immunochemical faecal occult blood test (iFOBT), to all Australians aged 50-74 years via the national postal service. The screening kit is self-administered at home and, if positive, a follow-up colonoscopy is recommended for diagnostic investigation. Prior to COVID-19, participation was approximately 44%, with 62% of individuals with a positive iFOBT reported as completing a colonoscopy.^5^ Using the Policy1-Bowel model we evaluated a 3-, 6-, or 12-month national-level pause, commencing 1 April 2020 during which time we assumed that no screening invitations or kits were sent or processed and all primary screening and diagnostic or surveillance colonoscopies associated with the NBCSP were halted. For modelling purposes, these periods were assumed to occur in 2020 – starting from April for 3- or 6-month pauses, and over the course of 2020 for the 12-month pause.

The status quo scenario is modelled based on no distruption to the NBCSP using observed participation rates (∼ 40%), with all rates from 2017 onwards extrapolated from the reported data for 2017, unless otherwise noted. Based on the observed data, the model assumes ∼ 70% of individuals with a positive iFOBT complete a diagnostic follow-up colonoscopy. Depending on the follow-up colonoscopy findings, individuals are referred to either return to the NBCSP for iFOBT screening after an interval of 4 years, or to repeat colonoscopic assessments (referred to as surveillance colonoscopies) after 1 to 5 years based on the 2011 guidelines recommendations. Combined, both follow-up and surveillance colonoscopies are referred to as NBCSP-related colonoscopies.

Outcomes have been estimated for the affected cohorts only i.e. among people aged 50-74 years eligible for screening in 2020 and 2021. This cohort comprises 7.1 million people; 3.5 million men and 3.6 million women.

Based on the GESA recommendations from 24th March 2020, most “elective” colonoscopies were suspended for most of March and April 2020 unless considered to be “urgent”. Recommendations suggested that colonoscopies for the investigation of a positive iFOBT be considered on a case-by-case basis only if the patient has not had a high-quality colonoscopy within the previous 4 years. The continuation of urgent colonoscopies was not incorporated in these results, nor was rescheduling of elective colonoscopies. We note that recommendations to recommence colonoscopy services were announced at the end of the April 2020 but have not been incorporated into these results.

Additional information on the Policy1-Bowel model can be found at www.policy1.org/models/bowel.

### Cervical screening

The National Cervical Screening Program (NSCP) is provided to women aged 25-74 years and usually involves a visit to primary care. In mid-March 2020, the NCSP was approximately 27 months into a transition from 2-yearly Pap testing to 5-yearly primary HPV screening, where a woman’s first HPV test was due two years after her previous Pap test.

As part of this evaluation, we updated modelled screening participation to reflect National Cancer Screening Register data (supplementary file 1B) indicating that 53.6% of women had already attended for their first HPV test, and therefore were not due to attend for routine screening again until at least December 2022. We evaluated 6-, 9-, or 12-month disruptions to screening participation in 2020, with participation during these periods being 50%, 75% and 90% lower, respectively, than would otherwise have been expected. We explored two recovery scenarios, one where women who missed screening in 2020 all attended in 2021, and another where women who missed screening in 2020 attended gradually over 2021-2022.

All modelled scenarios assume the NCSP transitioned from the pre-renewed NCSP (2-yearly cytology for women aged 18-69) to the renewed program (5-yearly primary HPV screening for women aged 25 to 70-74 years) at the beginning of 2018, including clinical management guidelines with women with detected abnormalities.^16-17^ The model incorporated recent policy changes that were expected to reduce the impact of COVID-19 disruptions, utilising unpublished NCSR data on the number of women who had received at least one primary HPV screening test and therefore were likely to be unaffected by the disruption (because they are not due for routine screening in 2020 or are already under surveillance). This meant that fewer women were expected to attend for a routine primary HPV test in Australia in 2020 than in 2019 or earlier years, and that women who would have attended in 2020 under the status quo were already overdue for screening (since it had been more than two years since their last Pap test), and as a result, a higher risk group.

Similar re-screening patterns as in the pre-renewed NCSP were assumed to apply until women have attended for their first Cervical Screening Test (CST) in the renewed NCSP (i.e., screening behaviour reflects that women are recommended to attend for their first CST two years after their last cytology test). These screening patterns for women’s first CST differ slightly from assumed adherence to the 2-yearly interval in the pre-renewed NCSP, in order to directly reflect NCSR data on observed behaviour from December 2017 onwards (see data sources for Policy1-cervix model at www.policy1.org/models/cervix). Re-screening patterns reflecting a recommended 5-yearly interval do not apply to women until after they have attended for their first CST. Assumptions for re-screening attendance (routine testing) and follow-up under all modelled scenarios are outlined in supplementary file 1C.

To calculate how many cancers were upstaged due to the disruption, we assumed that additional cancers that were diagnosed over 2021-2022 were diagnosed at the localised stage, and that any increase in the number of cancers diagnosed at the distant stage was due to cancers being upstaged from regional to distant. Other changes in the numbers of localised or regional cancers were assumed to be a result of upstaging from localised to regional.

### Reported outcomes

Outcomes reported from each modelled evaluation are shown in Table 1. Mortality estimates are not reported because it was not feasible to incorporate COVID disruptions to cancer treatment, which would also impact on cancer survival.

## Results

### Missed screens

For the scenarios modelled, the 3.7 million combined breast, bowel and cervical screening episodes expected under status quo in the 12 months starting 1 April 2020 would be reduced by 90% with a 12-month disruption and by 43% with a 6-month disruption (Table 2).

**Table 2:** Modelled screening episodes (number of screens and as a proportion of status quo screens) provided under each scenario over the period 1 April 2020 – 31 March 2021 for people in the target age range (eligible for screening during the affected period) and sex.

| Program (target sex and age range) | Disruption scenario by period |  |  |  |  | Observed number of tests as a proportion of status quo, Jan-Sept 2020 (by quarter and for whole period) (Q1, Q2, Q3, Q1-Q3) |
| --- | --- | --- | --- | --- | --- | --- |
|  | Status quo (comparator) | 12 months | 9 months | 6 months | 3 months |  |
| Breast (females, 50-74) | 973,019 (100%) | 0 (0%) | N/A | 298,113 (31%) | 518,680 (53%) | (98%, 44%, 105%, **82%**)<sup>α</sup> |
| Bowel (persons, 50-74) | 1,353,875 (100%) | 0 (0%)<sup>^</sup> | N/A | 680,259 (50%) | 1,016,915 (75%) | (54%, 92%, 97%, **81%**)<sup>β</sup> |
| Cervical (females, 25-74)*<sup>‡</sup> | 1,413,888 (100%) | 386,451 (27%) | 805,537 (57%) | 1,143,510 (81%) | N/A | (55%, 34%, 39%, **43%**)<sup>γ</sup> |
| **Combined - provided** | **3,740,782 (100%)** | **386,451 (10%)** | **N/A** | **2,121,882 (57%)** | **N/A** |  |
| **Combined - missed** | **0 (0%)** | **3,250,432 (90%)** | **N/A** | **1,612,032 (43%)** | **N/A** |  |

\* includes women attending for surveillance or other tests, not only routine primary screening tests. For this reason, the percentages do not correspond to the 25-95% reductions assumed among women attending for a routine cervical screening test.
<sup>^</sup>No screening provided during the reported period
<sup>‡</sup> For the cervical program scenario B is 9 months and scenario C is 6 months.
<sup>α</sup> Percentages derived from a comparison of screens for 2020 vs 2018, for women aged 50-74 years (AIHW table 1.2)<sup>25</sup>
<sup>β</sup> Percentages derived from a comparison of the number of kits returned for adults aged 50-74 years in 2020 vs 2019 (AIHW Table 3.2)<sup>25</sup>
<sup>γ</sup> Percentages derived from a comparison of the number of HPV tests conducted through the NCSP for women aged 25-74 years, 2020 vs 2019. (AIHW Table 2.2)<sup>25</sup>

### Breast cancer screening

#### Population cancer outcomes

For the two-year period 2020-2021, we estimate that screening pauses of any duration would lead to an overall reduction in population-level invasive breast cancer diagnosis for women aged 50-74 years, with a 12-month pause reducing cancer incidence by 9·4% (Table 3).

**Table 3:**
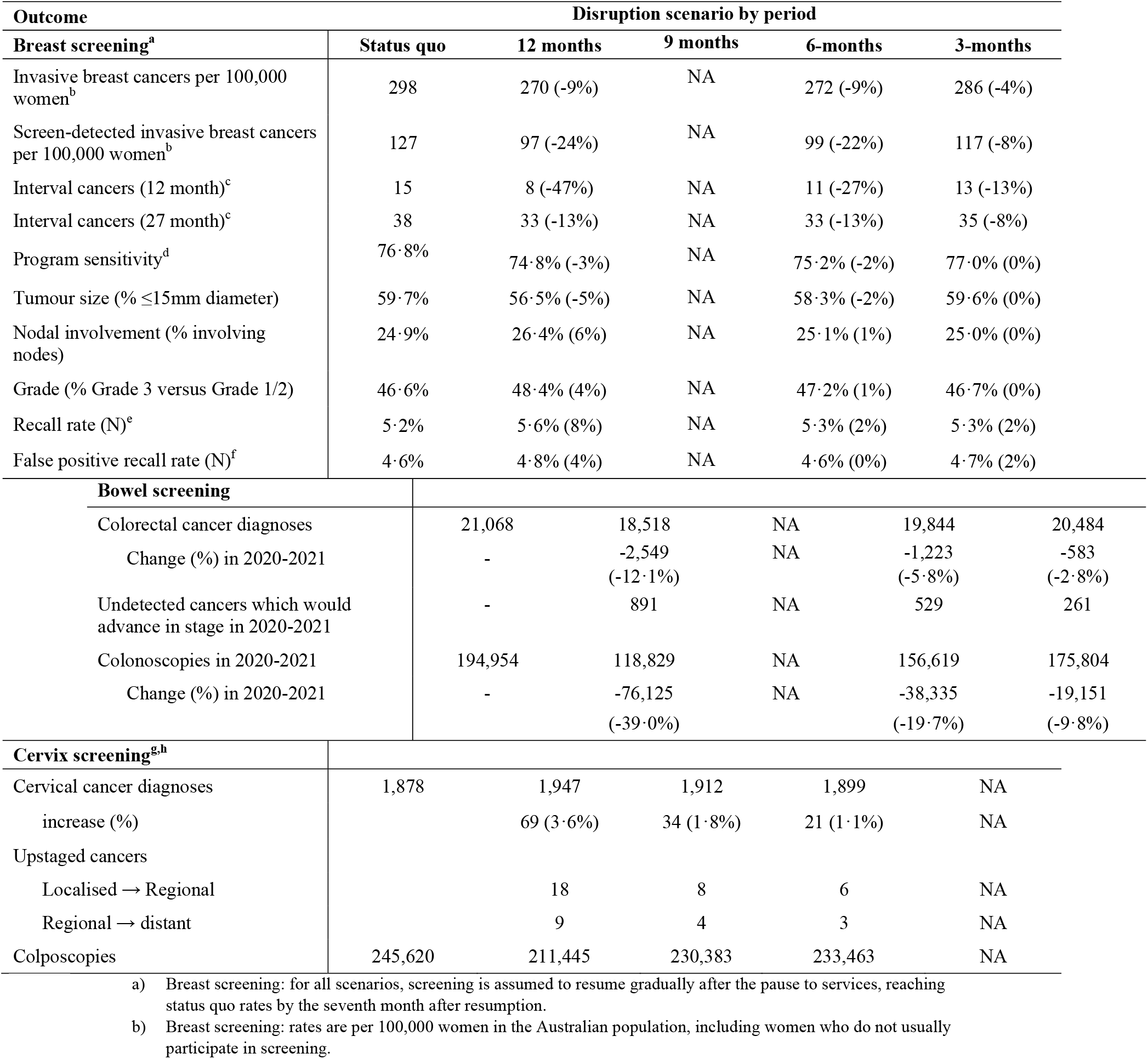
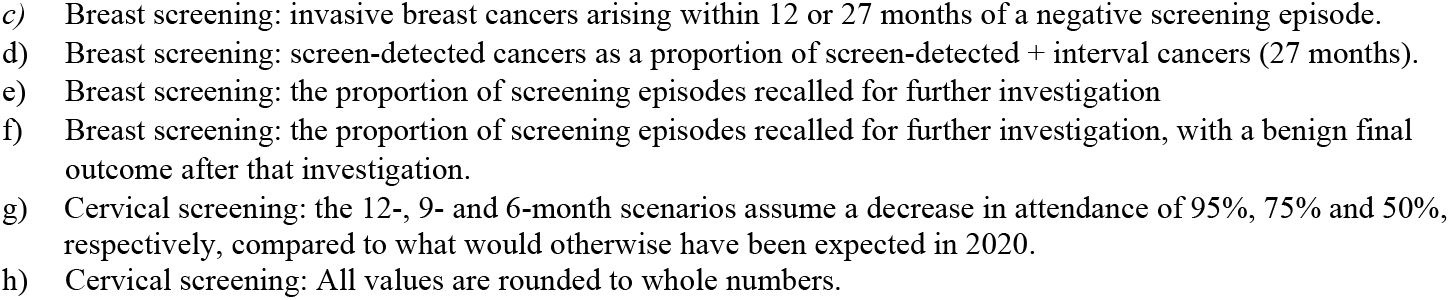
Selected estimated outcomes (2020-2021) for the evaluation of disruptions to breast, bowel and cervical cancer screening. Proportional changes compared to status quo are shown in brackets.

| Outcome | Disruption scenario by period |  |  |  |  |
| --- | --- | --- | --- | --- | --- |
| Breast screening<sup>a</sup> | Status quo | 12 months | 9 months | 6-months | 3-months |
| Invasive breast cancers per 100,000 women<sup>b</sup> | 298 | 270 (-9%) | NA | 272 (-9%) | 286 (-4%) |
| Screen-detected invasive breast cancers per 100,000 women<sup>b</sup> | 127 | 97 (-24%) | NA | 99 (-22%) | 117 (-8%) |
| Interval cancers (12 month)<sup>c</sup> | 15 | 8 (-47%) | NA | 11 (-27%) | 13 (-13%) |
| Interval cancers (27 month)<sup>c</sup> | 38 | 33 (-13%) | NA | 33 (-13%) | 35 (-8%) |
| Program sensitivity<sup>d</sup> | 76·8% | 74·8% (-3%) | NA | 75·2% (-2%) | 77·0% (0%) |
| Tumour size (% ≤15mm diameter) | 59·7% | 56·5% (-5%) | NA | 58·3% (-2%) | 59·6% (0%) |
| Nodal involvement (% involving nodes) | 24·9% | 26·4% (6%) | NA | 25·1% (1%) | 25·0% (0%) |
| Grade (% Grade 3 versus Grade 1/2) | 46·6% | 48·4% (4%) | NA | 47·2% (1%) | 46·7% (0%) |
| Recall rate (N)<sup>e</sup> | 5·2% | 5·6% (8%) | NA | 5·3% (2%) | 5·3% (2%) |
| False positive recall rate (N)<sup>f</sup> | 4·6% | 4·8% (4%) | NA | 4·6% (0%) | 4·7% (2%) |
| **Bowel screening** |  |  |  |  |  |
| Colorectal cancer diagnoses | 21,068 | 18,518 | NA | 19,844 | 20,484 |
| Change (%) in 2020-2021 | - | -2,549<br>(-12·1%) | NA | -1,223<br>(-5·8%) | -583<br>(-2·8%) |
| Undetected cancers which would advance in stage in 2020-2021 | - | 891 | NA | 529 | 261 |
| Colonoscopies in 2020-2021 | 194,954 | 118,829 | NA | 156,619 | 175,804 |
| Change (%) in 2020-2021 | - | -76,125<br>(-39·0%) | NA | -38,335<br>(-19·7%) | -19,151<br>(-9·8%) |
| **Cervix screening<sup>g,h</sup>** |  |  |  |  |  |
| Cervical cancer diagnoses | 1,878 | 1,947 | 1,912 | 1,899 | NA |
| increase (%) |  | 69 (3·6%) | 34 (1·8%) | 21 (1·1%) | NA |
| Upstaged cancers |  |  |  |  |  |
| Localised → Regional |  | 18 | 8 | 6 | NA |
| Regional → distant |  | 9 | 4 | 3 | NA |
| Colposcopies | 245,620 | 211,445 | 230,383 | 233,463 | NA |

a) Breast screening: for all scenarios, screening is assumed to resume gradually after the pause to services, reaching status quo rates by the seventh month after resumption.
b) Breast screening: rates are per 100,000 women in the Australian population, including women who do not usually participate in screening.
- c) Breast screening: invasive breast cancers arising within 12 or 27 months of a negative screening episode. - d) Breast screening: screen-detected cancers as a proportion of screen-detected + interval cancers (27 months). - e) Breast screening: the proportion of screening episodes recalled for further investigation - f) Breast screening: the proportion of screening episodes recalled for further investigation, with a benign final outcome after that investigation. - g) Cervical screening: the 12-, 9- and 6-month scenarios assume a decrease in attendance of 95%, 75% and 50%, respectively, compared to what would otherwise have been expected in 2020. - h) Cervical screening: All values are rounded to whole numbers.

All scenarios would lead to reduced population-level rates of screen-detected cancers and interval cancers, reduced program sensitivity, and more advanced tumour stage at diagnosis. Longer-term fluctuations in clinical and program outcomes are also expected; for example, with a 12-month pause we estimate a 10% difference in population-level invasive breast cancer diagnoses among women aged 50-74 between 2020-2021 (270 per 100,000 women) and 2022-2023 (296 per 100,000 women) (supplementary file 1D).

#### BreastScreen program outcomes

We assumed higher priority for women who missed scheduled screens during the pause, and report the estimated distribution of these women compared to other client groups (supplementary file 1E). Estimated screening intervals ranged from a median of 107 weeks for a 3-month pause through to 130 weeks for a 12-month pause, with a median of 154 weeks for women who missed screens during the pause (supplementary file 1E). Recall rates are predicted to fluctuate over time under various pause scenarios, ranging from 5·3% (3-month pause) to 5·6% (12-month pause) (Table 3), most likely due to an increasing proportion of first-round screening during recovery.

### Bowel cancer screening

#### Population cancer outcomes

All three scenarios illustrated a decrease in cancer diagnoses and cancers being diagnosed at a later stage in the screening cohort compared to the status quo. Across the modelled scenarios, colorectal cancer diagnoses decreased in 2020-2021 by 583 to 2,549 cases (2· 8-12·1% decrease). The modelled disruptions resulted in a stage shift, with up to 891 cases which would have been diagnosed but instead progressed to a later stage with lower survival. Considering both the reduction in colorectal cancer diagnoses in 2020-2021 and those which were diagnosed in 2021 rather than 2020, an estimated 759-3022 people would be affected by delays in diagnosis due to potential disruptions to routine screening in 2020-2021.

Due to the screening disruptions, fewer positive iFOBT would lead to fewer diagnostic follow-up colonoscopies, and additionally no surveillance colonoscopies would be conducted. The NBCSP- related colonoscopies that would not be conducted in 2020 due to the 3-, 6- and 12-month disruptions would be 19,151, 38,335 and 76,125 respectively.

This analysis found that a 12-month pause to screening would lead to an additional 7,140 colorectal cancer cases over 2020-2050. This is due to both changes in detection of both cancers and precancerous lesions during the 2020-2021 period, as well as the long-term effect on changes to screening participation and behaviours estimated to be attributable to the disruption.

Additional outcomes for diagnostic assessments and short-term adenoma outcomes are presented in supplementary file 1F.

### Cervical cancer screening

#### Population cancer outcomes

All three scenarios considered resulted in an increase in cancer diagnoses and cancers being diagnosed at a later stage among screening-age women compared to what would have been expected in the absence of any disruption. The increase in cancer cases over 2020-2022 ranged from 21-69 cases (1·1-3·6% increase). Most (57-63%) of the additional cancers were diagnosed among women 30-49 years. Women aged 30-39 years and 40-49 years were also the age groups where the percentage increase in cancers was largest (although still relatively small: 1·1-4·1% and 1·2-4·3% in women aged 30-39 and 40-49 years, respectively). The model predicted that disruptions to routine primary screening would lead to 6-18 cervical cancers being diagnosed at regional stage in 2021-2022, rather than as localised cancers in 2020; and 3-9 cervical cancers being diagnosed at distant stage in 2021-2022, rather than at regional stage in 2020 (Figure supplement 3). Considering both additional cervical cancers and those which were diagnosed at a later stage due to disruptions to cervical screening, an estimated 30–97 women would be affected by delays in diagnosis due to disruptions to routine screening in 2020 (Figure supplement 4 and 5). The longer-term impact of these additional and upstaged cancers on cervical cancer deaths are presented in supplementary file 1G and the impact of disruptions on the number of women expected to attend a cervical screening test are presented in supplementary file 1H (also see Figure supplement 6).

#### Colposcopy demand

Disruptions to primary screening in 2020 are estimated to result in 17,680–47,868 fewer women attending for a colposcopy in 2020 (17·4–47·2% reduction) as a result of their primary screening or triage test. The impact of the disruptions on expected colposcopy utilisation between 2020 and 2022 are presented in supplementary file 1I (also see Figure supplement 7).

#### Additional results

Additional results can also be found in the on-line reports produced as part of this project for the Australian Government, Department of Health.^19-21^

## Discussion

We present outcomes estimated in 2020 for a range of potential disruptions to national population cancer screening programs for breast, cervical and colorectal cancer, including population-level changes in cancer diagnoses and staging. For similar disruption scenarios, we estimate markedly different impacts for each program. We estimated that a 12-month screening disruption from early 2020 would in the short-term lead to reduced diagnoses of breast cancer (9·3% population-level reduction over 2020-2021) and colorectal cancer (up to 12·1% reduction over 2020-21) and increased cervical cancer diagnoses (up to 3·6% over 2020-2022), with upstaging expected for all three cancer types.

Findings were most marked for bowel screening, for which short-term upstaging and reduced cancer incidence was expected to lead to a significant number of additional colorectal cancer cases over the lifetime of the affected cohort. For breast screening, we estimated smaller but important reductions in cancer diagnoses at a population level over 2021 with fluctuating rates in subsequent years and upstaging of cancers at diagnosis (noting that estimated reductions in population-level interval cancer rates are counter-intuitive but an artefact of reduced screening participation, as interval cancers are defined with reference to screening events). For cervical screening, relative and absolute increases in cancer are expected to be less than for breast and colorectal cancer. This is likely due to COVID-19 impacts coinciding with lower participation under status quo because the program recently transitioned to a longer screening interval, along with the lower burden of disease due to prevention through both screening and widespread uptake of HPV vaccination (80% 3-dose coverage among females aged 14-15).^22^

### Strengths and limitations

We used established modelling platforms for breast, colorectal and HPV/cervical cancer natural history and screening in Australia, which were already well calibrated and validated to the Australian context using high quality published data, including detailed governmental screening reports. These models describe the Australian national age and sex distribution and the incidence and natural histories of breast, cervical and colorectal cancer, and can be used to simulate a range of scenarios for cancer screening. Prior investment into these modelling platforms facilitated the rapid response modelling required in the face of a crisis; the evaluations reported highlight the importance of ongoing investment into comprehensive, well-calibrated and validated population predictive models that can be quickly harnessed for critical decision making as needed for disruptions such as the COVID-19 pandemic and other potential disruptions due to major social, public health or natural disasters as are increasingly common due to climate change.^23^

This evaluation included detailed reporting of outcomes for different sub-groups where this was requested by key stakeholders (for example, breast screening outcomes according to whether clients were directly or indirectly impacted by screening disruptions).

Each modelled evaluation drew on the most contemporary data available, employing projections as required. In addition, the cervical screening model incorporated the effects of recent policy changes that were expected to reduce the impact of COVID-19 disruptions, drawing on unpublished data from the NCSR.

Scenarios were by necessity defined in simple terms. For example, we modelled single, continuous periods of national disruption but, in practice, screening participation would likely fluctuate by time and place in line with local outbreaks and pandemic control measures. By deliberately focusing on a high-level effect – a wide range of possible effects on attendance for routine screening – the analysis was agnostic to the cause, and therefore this could reflect any or many of a wide range of possible factors, including changes in behaviour and access to screening, reduced capacity of services involved in providing screening, or reduced saliency or delivery delays of reminder letters or home-delivered testing kits.

Modelled estimates will likely include some biases. We do not account for changes in diagnostic and treatment services due to COVID-19 which have since been observed.^24^ For example, Medicare Benefits Schedule claims data from 2020 indicates a 4% reduction in breast diagnostic services and 6% reduction in breast cancer related surgical procedures, and for colorectal cancer a 13% reduction in diagnostic services (mostly colonoscopies) and 1% reduced surgical procedures. For cervical cancer screening, estimates of additional and upstaged cervical cancer diagnoses potentially underestimate the overall impact of the pandemic on cervical cancer diagnoses.

### Subsequent observed outcomes

This modelled evaluation was conceived during the first wave of COVID-19 restrictions, and in the context of the need to provide safe health services including those for Australia’s established population screening programs for breast, cervical and colorectal cancer. At the time, and with the current analysis providing background estimates of potential impacts, the Australian cervical and bowel screening programs continued to operate continuously through 2020 (and since); BreastScreen services paused nationally from March to late April/early May 2020, resuming thereafter.

Australian cancer screening programs reported reduced and fluctuating screening tests for all programs over the period January-September 2020 (Figure 1 and Figure 2).^25-26^

**Figure 1:**
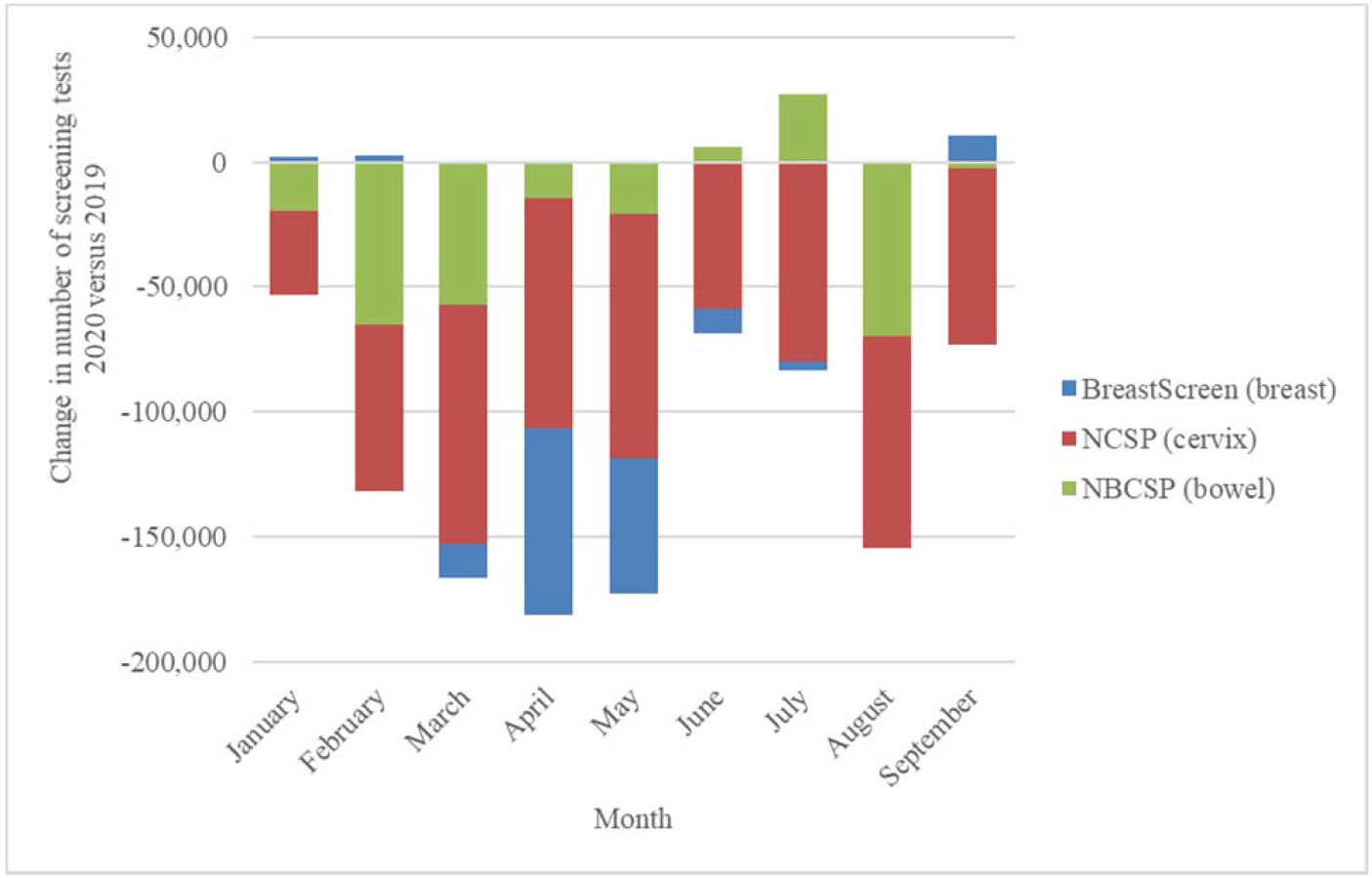
National change in number of screening tests by month, 2020 versus 2019, for the breast, cervix and bowel screening programs. Derived from AIHW 2020 report – cancer screening and covid19 in Australia. ^25^ Note: Cervical screening test volumes were anticipated to be lower in 2020 than in 2019 due to the extension from a 2-year to a 5-year screening interval.^26^

**Figure 1.**
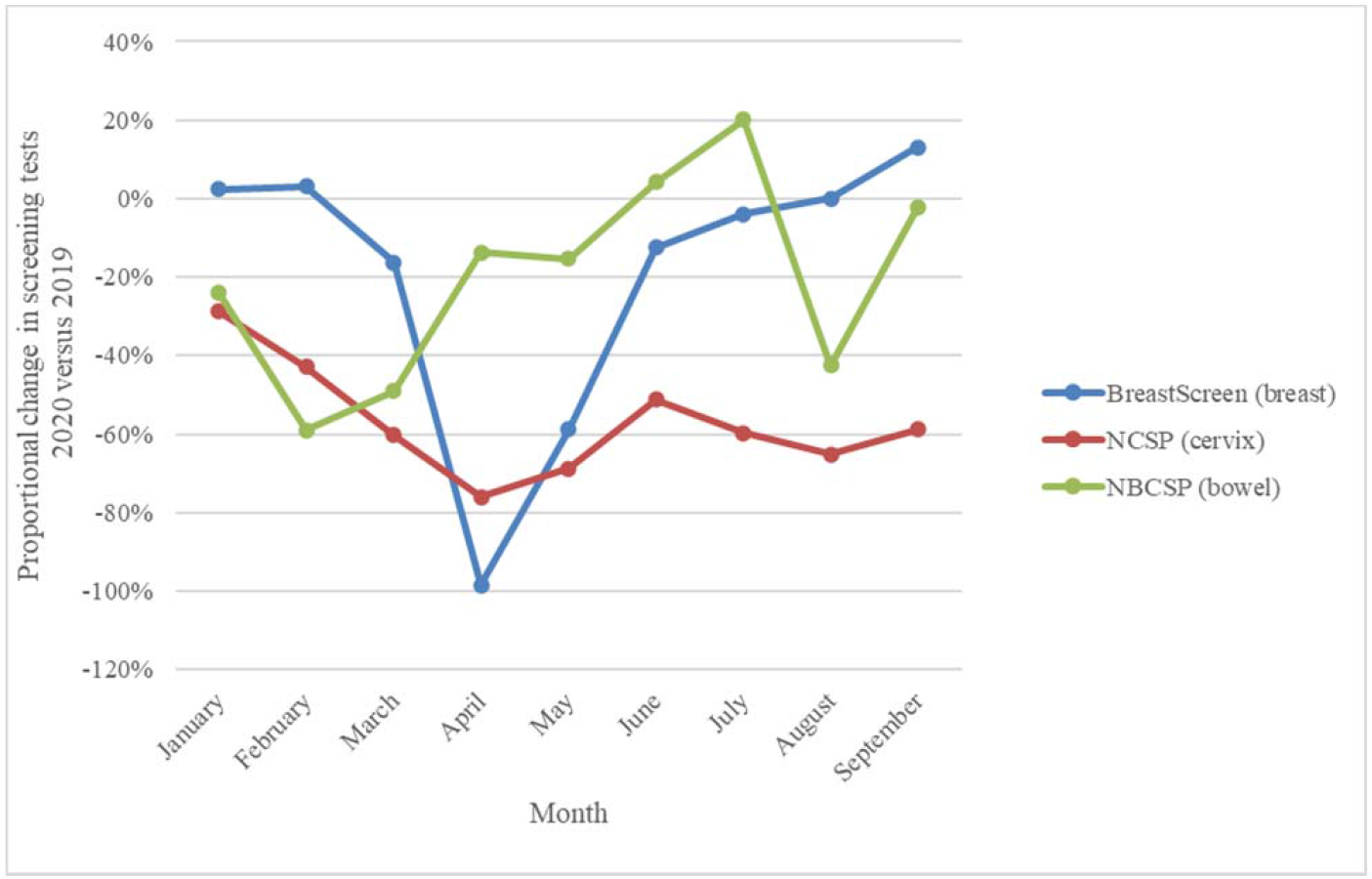
National proportional change in number of screening tests by month, 2020 versus 2019, for the breast, cervix and bowel screening programs. Derived from AIHW 2020 report – cancer screening and covid19 in Australia.^25^ Note: Cervical screening test volumes were anticipated to be lower in 2020 than in 2019 due to the extension from a 2-year to a 5-year screening interval. ^26^

Overall, screening tests across the three programs dropped by 34%, with over one million (1,035,710) fewer breast, bowel and cervix tests in age-targeted Australian populations from January to September 2020 compared to January to September 2019.^25^ Our model-estimated reductions in screening tests in the first year of disruption overlapped with observed annual and quarterly rates in program-specific screening tests (Table 2).

Observed screening reductions were driven by a range of factors such as screening service closure followed by reduced throughput capacity for breast screening, reduced primary care visits (cervical screening), as well individuals likely de-prioritising screening in the context of the pandemic.^27^ Screening test rates varied markedly between states and territories during the pandemic,^25^ consistent with localised, often state-and-territory based controls including stay-at-home orders, border closures, quarantine programs and other physical and social distancing measures. Our evaluation of a range of scenarios indicates the expected nature, scale and timeframe of key outcomes for each screening program, providing valuable insights relevant to both national and more localised disruptions.

### Future insights

Our predicted clinical implications of reduced participation will not be confirmed for some time, however, early signs suggest an observed reduction in cancer diagnosis in 2020. ^28^ While COVID-19 vaccination coverage is high in Australia (two dose: >95% nationally for people >16 years old; three dose:71% >16 years) ^29,30^ social distancing measures are a continuing requirement and the redeployment of health services may be required in future if new variants of concern with higher transmissibility and/or virulency take over. As observed data on COVID-impacted screening behaviour for specific sub-populations becomes available, further modelling can help estimate the impact of screening disruptions for different population sub-groups, noting that Australian screening data is regularly reported by age group, sex (for bowel cancer screening), and various socio-economic indicators. Each screening program is subject to detailed monitoring and evaluation, working to a range of performance indicators. These indicators are expected to be impacted by service disruptions, in a variety of ways. For example, breast screening interval cancer rates (a key BreastScreen performance indicator) are predicted to vary; this is logical given that the likelihood of an interval cancer is expected to depend on the time since the screen prior to the interval cancer reference screen, screening round, and age, all of which vary for the scenarios evaluated. These changes would impact usual quality assurance monitoring of the program and, potentially, community perception of program performance. Therefore, one of the implications of the current evaluation is to focus future strategies for communication around the importance of screening programs and the need for ongoing high participation as they play a key role in maintaining community confidence in screening.

We quantified resource requirements for programs and related health services in the face of disruption, with requirements for breast cancer screening assessments, colonoscopies and colposcopies affected to varying degrees. Variations in breast and colorectal cancer incidence are expected to have a significant flow-on effect on the demand for treatment services, compounded by a changing case-mix with shifts to later-stage diagnoses to varying degrees over time.

### Implications

Collectively, these findings illustrate that maintaining screening participation is critical to reducing the burden of cancer at a population level. We enumerate the extent to which COVID-19 disruptions are likely to impact short- and long-term cancer outcomes, resource requirements and usual indicators used in program monitoring and evaluation, providing critical evidence to guide cancer screening programs as they continue to adapt to the ongoing impacts of the COVID-19 pandemic, and as they prepare for other major disruptions that may arise in the future.^28^

The estimated impacts described here may be reduced if screening programs sustain or revisit planned improvements that commenced prior to the pandemic, some of which may also directly assist program recovery. For example, from 1 July 2022, all women now have the option to use self-collection and this could facilitate continuation or recovery of cervical screening programs.^30^

As programs work to bring screening participants back on schedule, well-planned reactive risk-based approaches may help direct limited services to those who will benefit the most. For HPV screening, this could mean more refinements to triaging and surveillance of screen-positive women, and optimising screening for women protected by HPV vaccination. For breast screening, this could involve prioritising women at higher risk of breast cancer, such as women usually offered annual screening and women assessed as higher risk using routine risk prediction models incorporating breast density.^31^ For bowel screening, risk-stratified prioritisation of people could be rapidly implemented by modifying the faecal occult blood threshold and extending the period of time over which people are asked to complete their missed screens.^32^ Future improvements could also consider starting screening at an earlier age for Aboriginal and Torres Strait Islander peoples who are at higher risk of colorectal cancer.^33^ For all screening programs, individuals who missed screening during the disruption should be encouraged to return to screening as soon as it is safe to do so. For bowel screening, previously published modelling estimated that this “catch-up” screening could almost ameliorate the impact of a disruption.^12^ As such, one implication of the current evaluation is that screening programs have the potential to continue to improve outcomes and therefore “build back better” in the future.

## Data Availability

Supporting information contained in the supplementary materials provide details on microsimulation model parameters, calibration to epidemiologic data, and calibration approach in line with good modelling practice. The current manuscript is a computational study involving modelling rather than direct analysis of primary datasets.

## Acknowledgments

Australian Government Department of Health commissioned Cancer Council NSW to develop and conduct the modelled evaluations reported in this manuscript.

## Data availability statement

Supporting information on data sources informing each model can be found on-line (www.policy1.org). The current manuscript is a computational study involving modelling rather than direct analysis of primary datasets. Furthermore, no new datasets have been created.

## Code availability

Codes for the Policy1 models have been developed over decades, are proprietary property, and cannot be provided by the authors at this time; however, the Daffodil Centre is working towards providing transparent and reproducible modelling code in future. Access to current code is possible only through supervised training at the Daffodil Centre after submission and approval of proposal by interested researcher.

---

**Supplementary Figure 1-.**
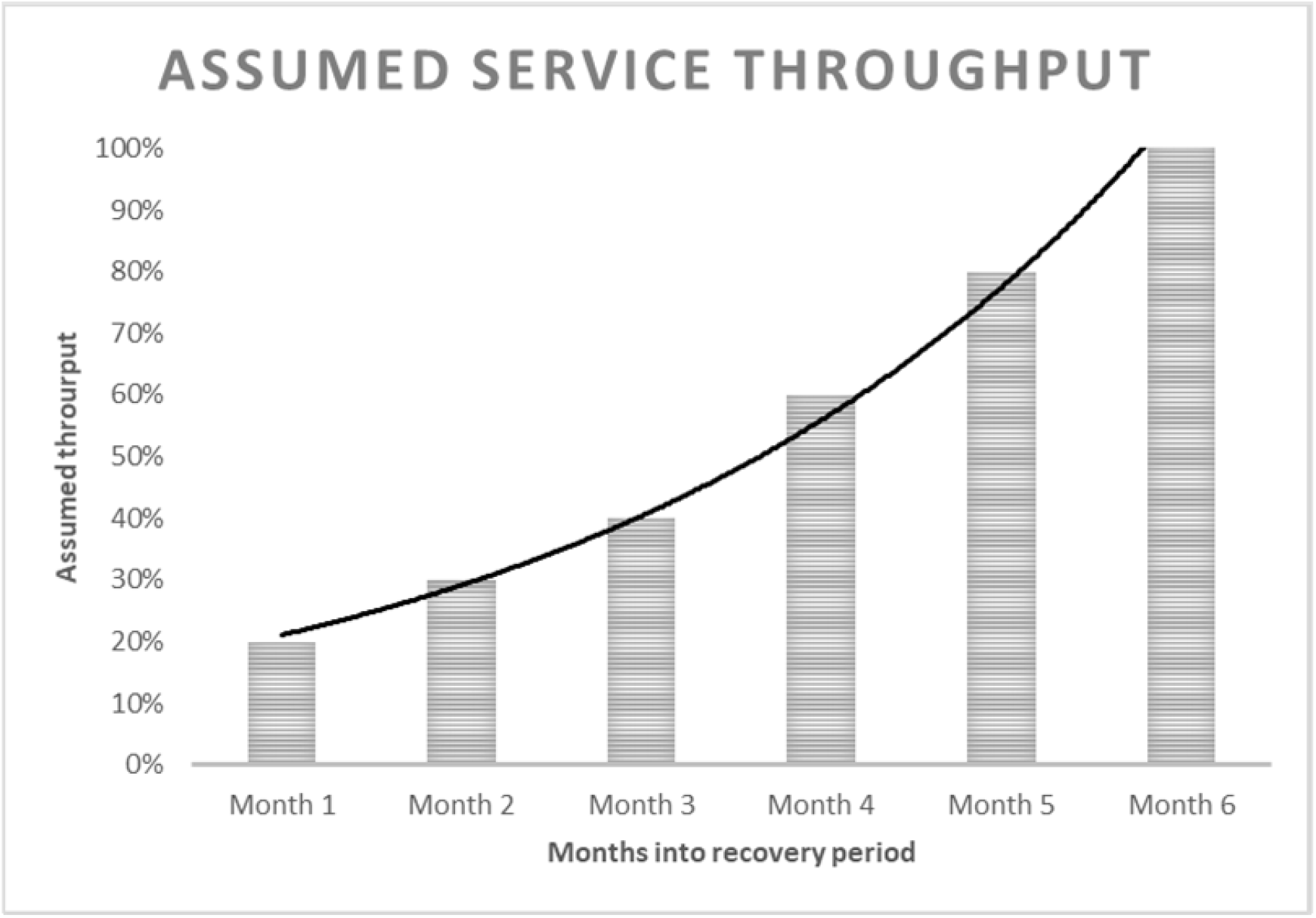
Gradual restoration of BreastScreen Australia service throughput as modelled during the first 6 months of the recovery period.

**Supplementary Figure 2-.**
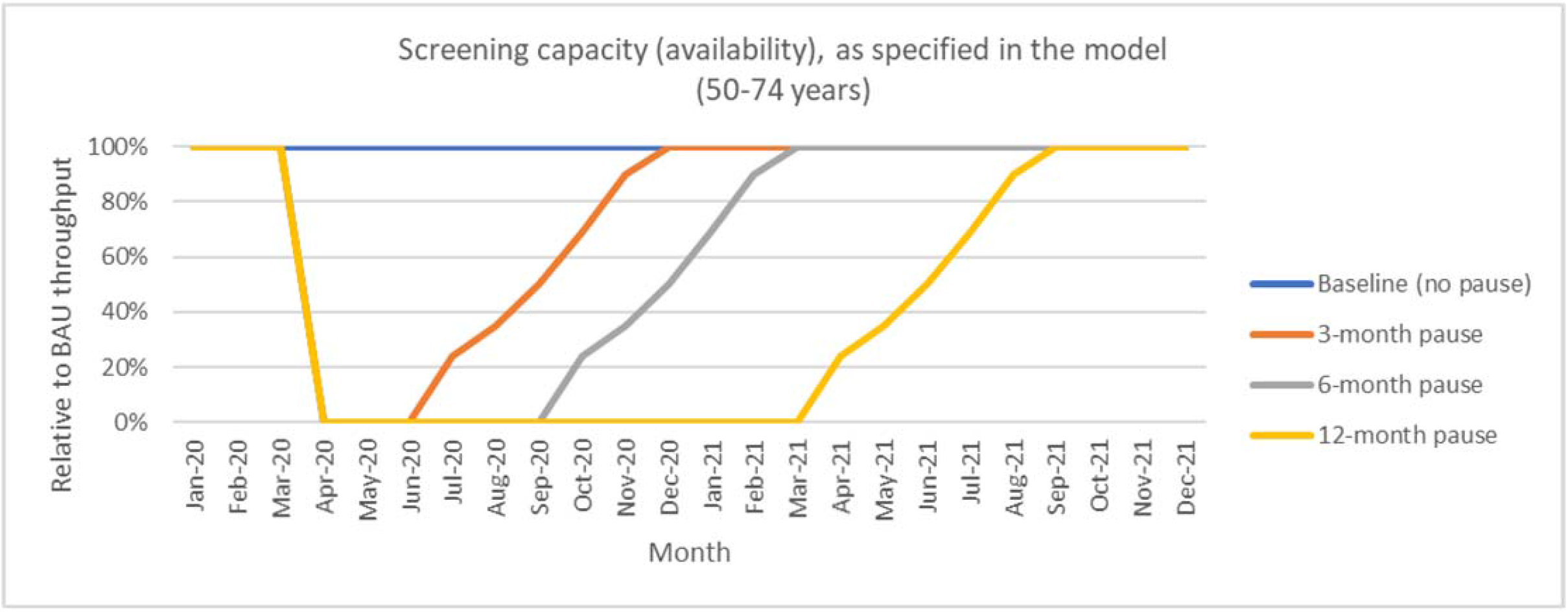
The combined effect of pause periods and gradual restoration of service throughput, as specified in *the model*.

**Supplementary Figure 3:**
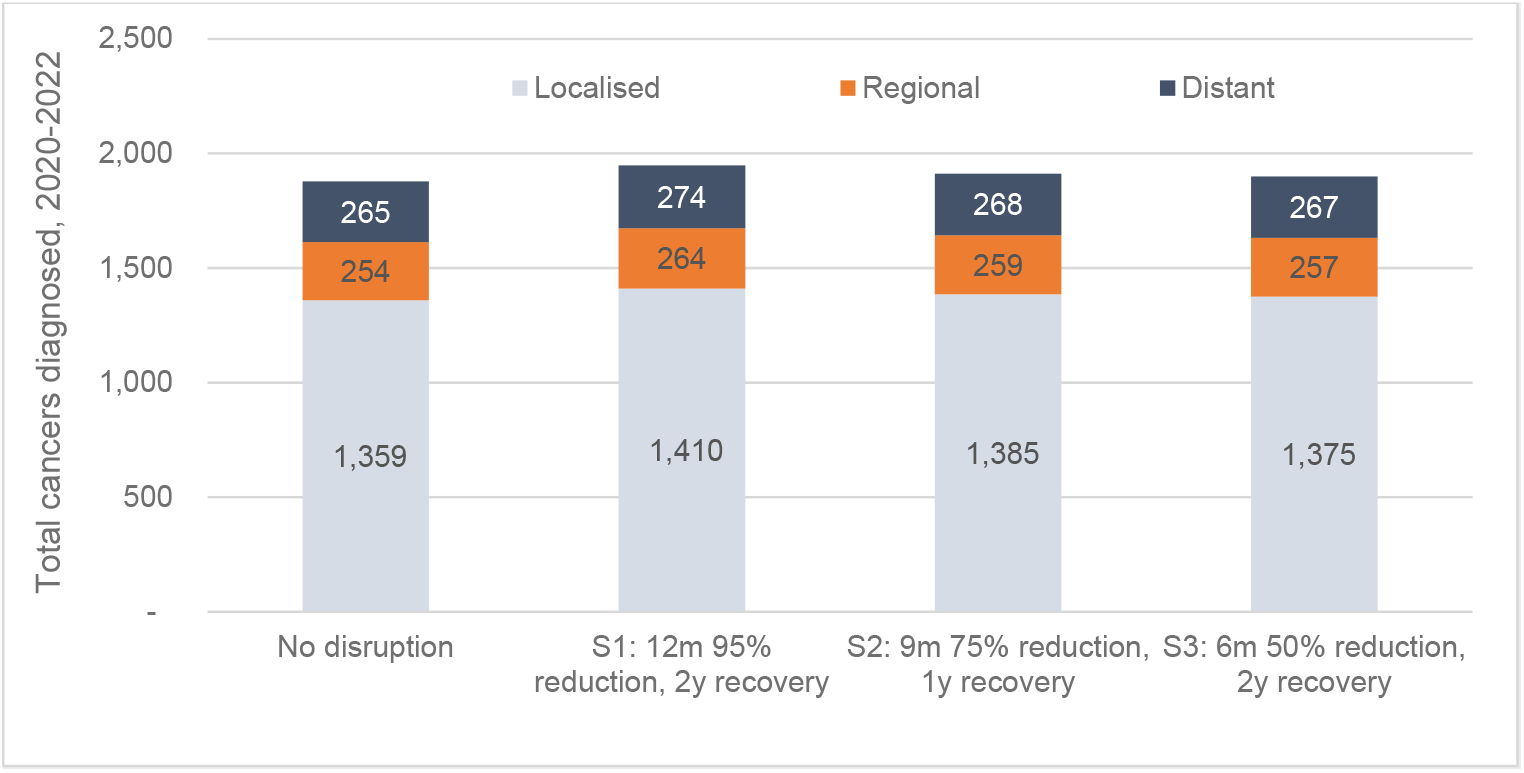
Total cervical cancers diagnosed among women aged 25-74 years in 2020-2022 under varying scenarios for disruptions to routine screening in 2020, by stage at diagnosis.

**Supplementary Figure 4:**
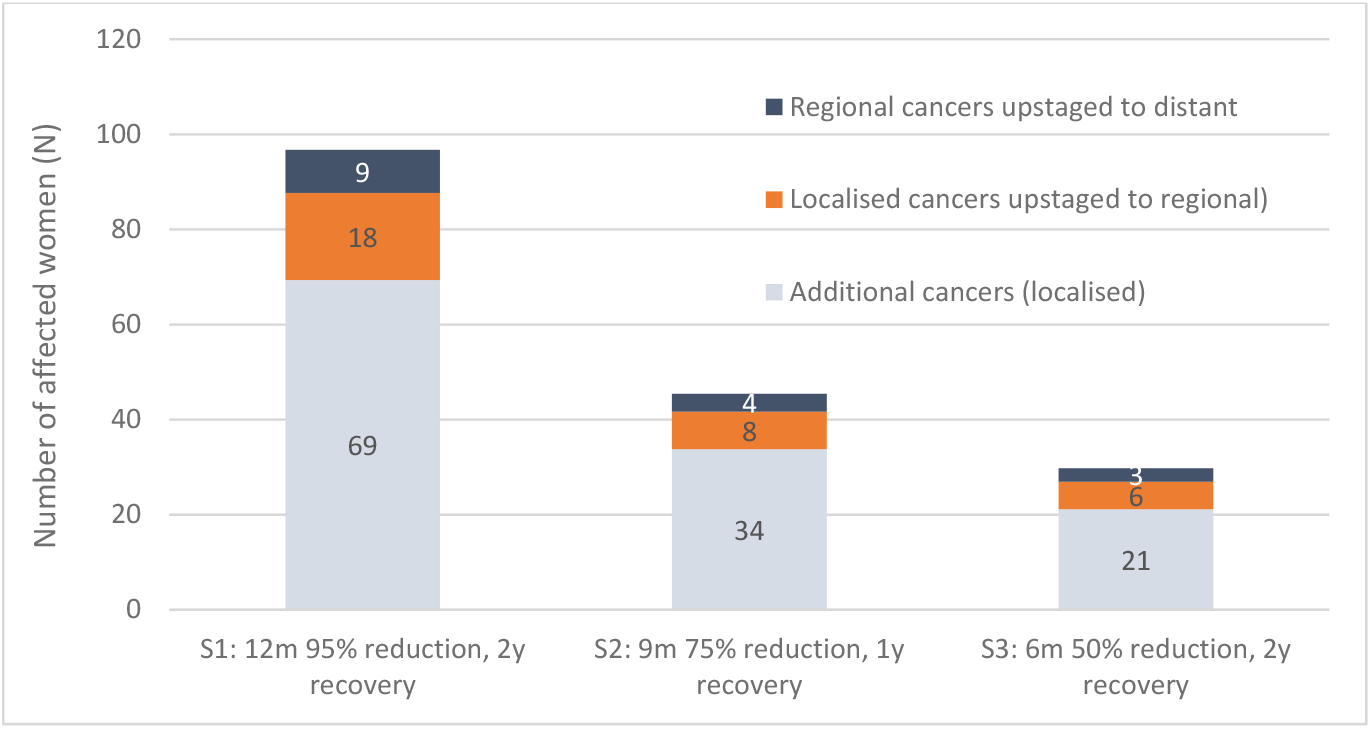
Number of women affected by delayed cancer diagnosis under varying scenarios for disruptions to routine screening in 2020. Upstaged regional cancers that progressed to distant were calculated by assuming that the difference in numbers of diagnosed distant cancers is due to upstaging from regional only. Upstaged localised cancers that progressed to regional were calculated by assuming that the number of upstaged cancers is equal to the difference in numbers of diagnosed regional cancers, corrected for the number of regional cancers which were upstaged to distant. Some differences between Figures 3 and 4 are due to rounding cases to whole numbers.

**Supplementary Figure 5:**
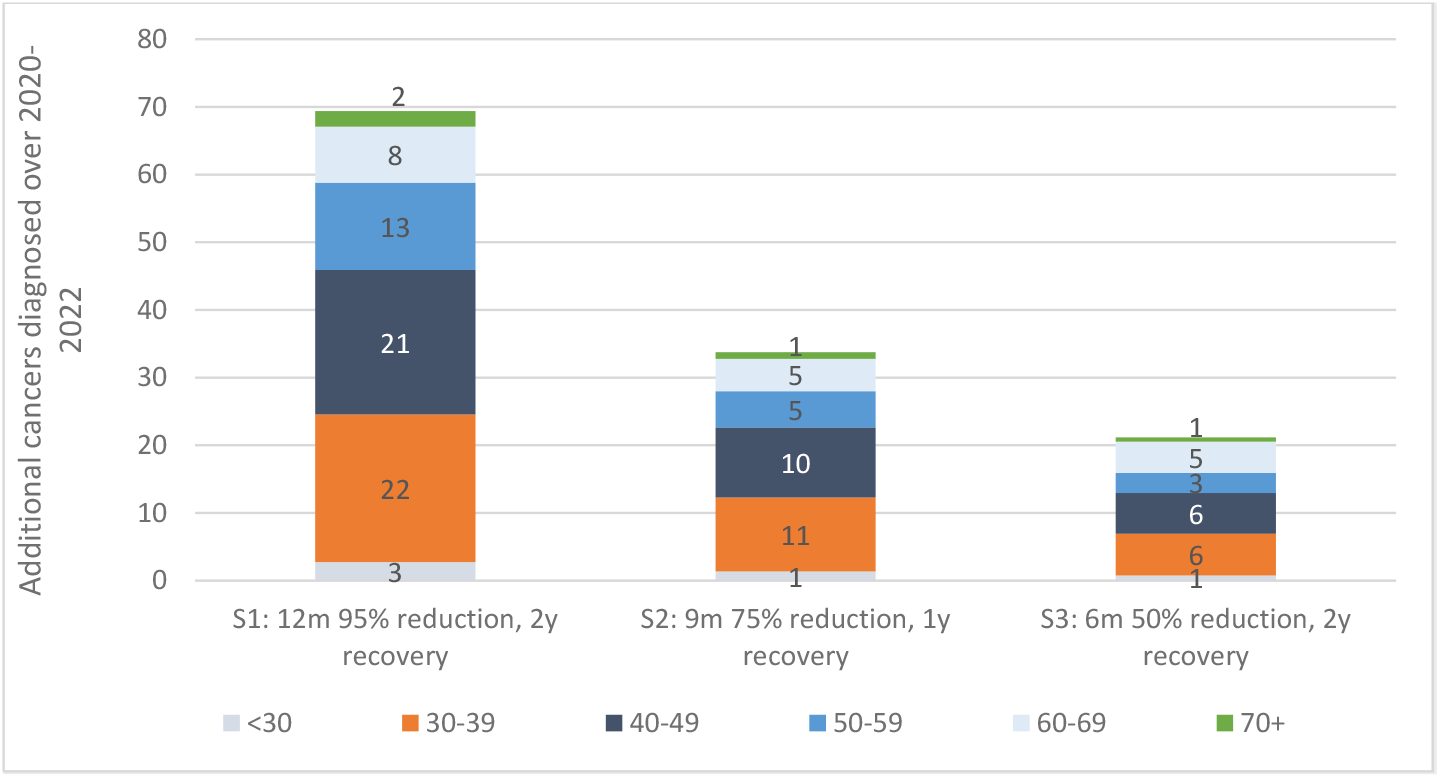
Additional cervical cancers diagnosed among women aged 25-74 years in 2020-2022 under varying scenarios for disruptions to routine screening in 2020, by age at diagnosis. Note: women aged <30 years in 2020 were offered HPV vaccine when aged <17 years; women aged 30-39 years in 2020 were offered HPV vaccination when aged 18-26 years

**Supplementary Figure 6-.**
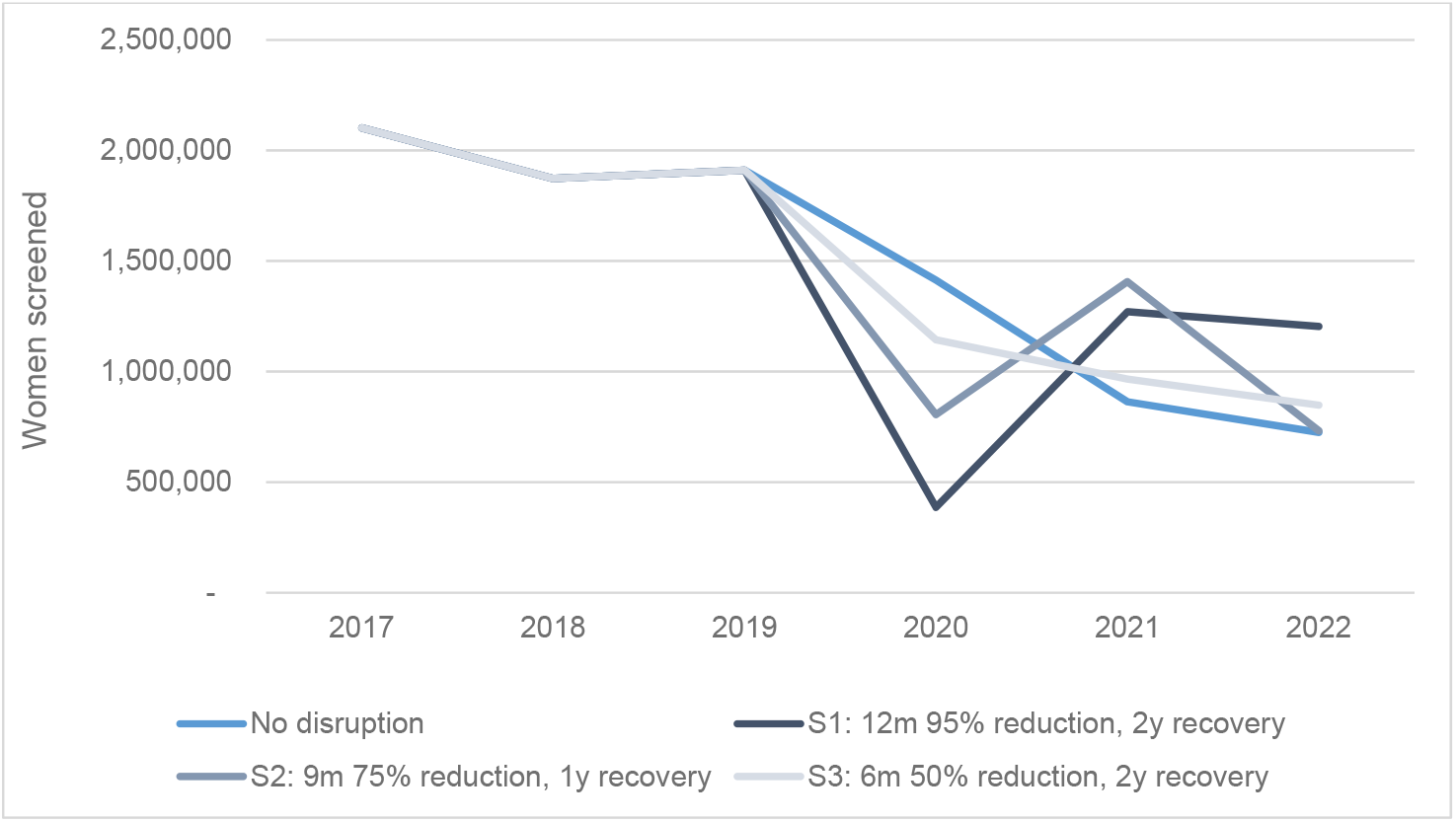
Predicted number of women aged 25-74 years attending for cervical screening under varying disruption scenarios, by year. Note: Data presented for each year are the total number in that year (existing model structure does not allow for predictions at the level of months)

**Supplementary Figure 7:**
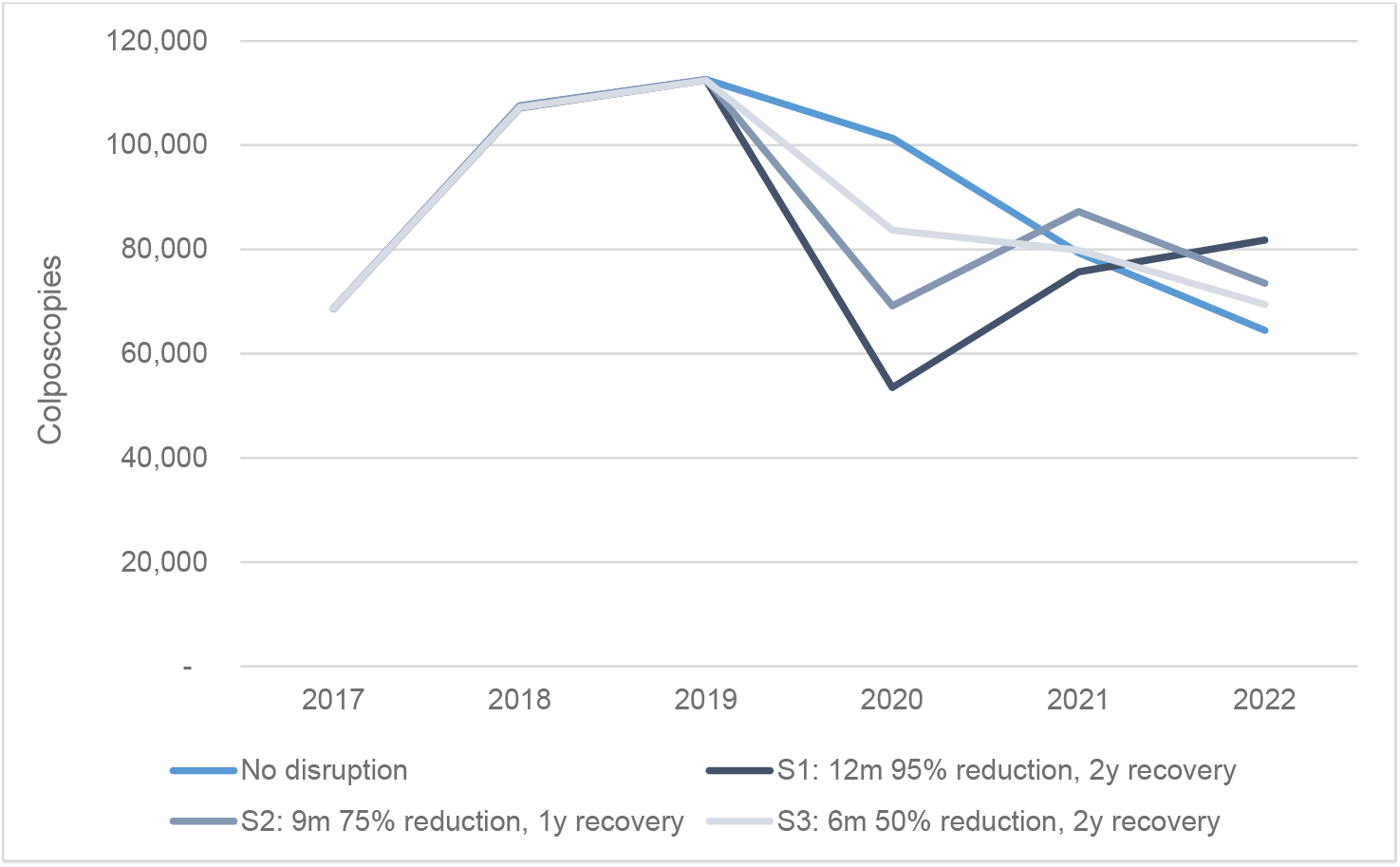
Predicted number of colposcopies among women aged 25-74 years under varying disruption scenarios, by year. Note: Data presented for each year are the total number in that year (existing model structure does not allow for predictions at the level of months). Note that the number of colposcopies that were anticipated in each of 2020, 2021, and 2022 was expected to differ year to year even in the no disruption scenario as a result of the transition from a 2-year to a 5-year recommended screening interval. This will remain the case in the context of a disruption, but the pattern will vary.

